# Comparative assessment of combined concentration and extraction methods for Influenza A and B virus detection in wastewater

**DOI:** 10.1101/2025.02.21.25322585

**Authors:** Till Fretschner, Edgar Elon Zeisler, Steffi Scheller, Andreas Aurenz, Beate Schneider, Marcus Lukas, Ulrike Braun, Timo Greiner, Jakob Schumacher, Hans-Christoph Selinka, Birgit Walther, René Kallies

**Author notes:** These authors share last authorship (Birgit Walther, René Kallies).

## Abstract

Influenza, caused by Influenza A and B viruses, represents a significant global health burden due to recurrent seasonal epidemics and the risk of pandemics. To gauge the large volume of seasonal influenza cases, it may be helpful to complement classical surveillance systems with additional approaches such as wastewater-based epidemiology (WBE), which can aid in the early trend assessment of seasonal epidemics. WBE has emerged as a promising tool for population-level surveillance, enabling the detection of viral nucleic acids in wastewater and offering unique advantages over individual-based surveillance. This study evaluates the performance of six different combinations of virus concentration and RNA extraction methods for the detection of Influenza A and B viruses in wastewater. Composite samples from four wastewater treatment plants in North Germany were analyzed using precipitation, filtration and automated extraction protocols. Method performance was evaluated by quantitative recovery of viral RNA and a spike-and-recovery experiment. The combination of PureYield™ filtration and Maxwell® RSC extraction (PYC_EX1) consistently demonstrated the highest recovery rates for both Influenza virus A and B, achieving recovery efficiencies of up to 80.4% and 72.3%, respectively. This method also enabled reliable detection of low viral loads, which is critical for an early detection of rising incidence. Our findings underscore the importance of rigorous method evaluation to optimize WBE for influenza surveillance. By providing robust, sensitive and reproducible protocols, this study highlights the potential of WBE to improve public health preparedness, enables timely interventions and reduces the spread of influenza viruses within communities.

## 1 Introduction

Influenza, a highly contagious respiratory illness, poses a significant global health threat, causing recurring seasonal epidemics associated with substantial morbidity and mortality worldwide (Harrington et al., 2021). The primary culprits are Influenza A (IAV) and B (IBV) viruses, which drive most seasonal outbreaks. Other types, such as Influenza C and D viruses, also exist and can cause infections, though their public health impact is less pronounced (White et al., 2016; Sederdahl and Williams, 2020; Hammond et al., 2022). In 2019, the World Health Organization (WHO) highlighted the threat of another severe influenza pandemic as one of the top ten global health concerns, underscoring the urgent need for preparedness (WHO, 2019). Surveillance systems play a crucial role in controlling the spread of Influenza by providing timely and accurate data to public health (PH) authorities (Hammond, Kim, and Vandemaele 2022; WHO 2013). However, traditional surveillance methods, such as passive, active, and sentinel systems, primarily capture data from individuals seeking medical care. As a result, they may underestimate the true burden of infection within the community, particularly in individuals with mild or asymptomatic courses who continue to contribute to viral transmission (Montgomery et al., 2024).

Wastewater-based epidemiology (WBE) has gained increasing attention as an innovative tool for public health surveillance, particularly during the COVID-19 pandemic (Kramarsky-Winter et al., 2023; Carmo dos Santos et al., 2024). By detecting viral nucleic acids in wastewater, WBE provides a more comprehensive picture of community-level infections, independent of healthcare-seeking behavior. This approach provides a valuable complement to traditional individual-based surveillance methods, (Kilaru et al., 2023; Li et al., 2024).

Influenza viruses, particularly Influenza virus A and B, are among the most important causes of respiratory infections, making them desirable targets for WBE. However, despite its success with viruses like SARS-CoV-2, WBE for influenza virus remains underdeveloped (Grassly et al., 2025).The challenges include the detection of relatively low viral loads in wastewater and the need for highly sensitive and specific methods to extract and quantify influenza viral RNA (Cheshomi et al., 2024). Implementing WBE for influenza virus requires overcoming these technical hurdles, but successful detection could improve Influenza surveillance by providing earlier warnings of outbreaks and better capturing the dynamics of virus spread within communities.

The reliability of WBE for detecting influenza virus depends heavily on the quality of the methods used for virus concentration and RNA extraction. Influenza virus RNA in wastewater is often present in low concentrations, especially outside of peak transmission seasons, making sensitive detection methods crucial (Boehm et al., 2023a; Maida et al., 2024). Effective testing requires a combination of virus concentration and nucleic acid extraction techniques that are robust enough to work with the complex and variable matrix of wastewater. These methods must be optimized not only for sensitivity and specificity but also for practical considerations such as costs, processing time, and equipment availability. Factors such as sample volume, the potential presence of inhibitors in wastewater, and the viral recovery rate significantly influence the success of the detection process (Zafeiriadou et al., 2024).

In this study, we evaluated various combinations of concentration and extraction methods to identify the optimal approach for detecting Influenza A and B virus RNA in wastewater. As part of the AMELAG (German: *Abwassermonitoring zur epidemiologischen Lagebewertung*) project, one of Germany’s largest wastewater monitoring initiatives (Marquar et al., 2024; Saravia et al., 2024), we conducted a series of experiments to evaluate specific virus concentration and RNA extraction methods for SARS-CoV-2 and surrogate markers. These methods were selected based on their proven effectiveness within AMELAG but do not necessarily represent the variety of methods for influenza virus detection in WBE. In the present paper, our goal was to identify the combination of methods that provide the highest recovery rates for Influenza A and B virus RNA in wastewater samples.

Initially, 24-hour composite samples from multiple wastewater treatment plants (WWTP) were collected and processed using six different method combinations. These methods included both precipitation and filtration-based concentration techniques, paired with automated nucleic acid extraction protocols. Performances of various method combinations were evaluated with regard to detection of Influenza A and B RNA, with Pepper Mild Mottle Virus (PMMoV) serving as an internal process control. Next, we conducted a spike-and-recovery experiment to assess the recovery efficiency of the best-performing method combination. Inactivated Influenza A and B viruses were spiked into raw wastewater at three different concentration levels, and the recovery rates were determined by comparing the detected viral loads to the known input concentrations.

Our study highlights the critical importance of thorough method evaluation to ensure reliable detection and quantification of influenza viruses in wastewater. Simultaneous improvements of both concentration and extraction techniques can avoid misinterpretation of results and allows provision of accurate data for public health authorities, thereby enhancing their ability to make informed decisions for the prevention and control of Influenza outbreaks.

## 2 Material and Methods

### 2.1 Wastewater Collection

A total of 16 raw wastewater samples were collected from four different wastewater treatment plants (WWTP) in Northern Germany during the seasonal Influenza, calendar week (CW) 46/2023 to 21/2024. Twenty-four-hour composite samples of grit removed wastewater were transported at 4°C to the Microbiological Risks Laboratory of the German Environment Agency on the same day. For the spike-and-recovery experiment, three liters of a 2-hour composite sample were collected and further processed (see below for detailed description). After homogenization for 15 minutes, 50 ml aliquots were centrifuged at 250 x g at 4°C for 15 minutes to remove solid matter.

### 2.1 Concentration Methods and Viral RNA Extraction

Three different concentration methods were performed. Forty ml supernatant of each sample were precipitated with Polyethylene glycol 8000 (Carl Roth, Karlsruhe, Germany), subsequently abbreviated as PEG, centrifugal filtration with Centricon 70-Plus filter cartridges (Merck, Darmstadt, Germany), abbreviated as CEN and PureYield™ Filtration using a silica membrane with Vac-Man® Laboratory Vacuum Manifold (Promega, Walldorf, Germany), abbreviated as PYC. RNA was extracted using either the Maxwell® RSC Enviro Total Nucleic Acid Kit (Promega, Walldorf, Germany) on a Maxwell® RSC automated instrument (Promega), abbreviated as EX1 or the innuPREP AniPath DNA/RNA Kit (Analytik Jena, Jena, Germany) on an InnuPure® C16 touch (Analytik Jena, Jena, Germany) system, abbreviated as EX2, following the manufacturer’s instructions. In contrast to the manufacturer’s protocol RNA was eluted in 100 µl RNase-free water. A summary of the different concentration and nucleic acid extraction methods is given in table 1.

**Table 1:** Combinations of concentration and extraction methods.

| Combination | Concentration Method | Extraction Method |
| --- | --- | --- |
| PEG/EX1 | Polyethylene Glycol (PEG) 8000 | Maxwell® RSC Instrument |
| CEN/EX1 | Centricon™ Plus-70 Filtration | Maxwell® RSC Instrument |
| PYC/EX1 | PureYield™ Filtration | Maxwell® RSC Instrument |
| PEG/EX2 | Polyethylene Glycol (PEG) 8000 | InnuPure® C16 touch |
| CEN/EX2 | Centricon™ Plus-70 Filtration | InnuPure® C16 touch |
| PYC/EX2 | PureYield™ Filtration | InnuPure® C16 touch |

### 2.2 Reverse Transcription Droplet Digital PCR (RT-ddPCR)

The RT-ddPCR was performed using the QX600 AutoDG Droplet Digital PCR System (Bio-Rad Laboratories, Hercules, Ca, USA) with the QX Manager Standard Edition v2.1.0 software, following a previously published protocol (Markt et al., 2023). The One-Step RT-ddPCR Advanced Kit for Probes (Bio-Rad Laboratories) was used with specific primers, probes, and positive controls for Influenza viruses A and B, as described more detailed in the Supplementary Information (LINK SI Table 1). Each RT-ddPCR run included both, a negative and positive control for each target virus. The forward and reverse primers were used at a final concentration of 1 µM, probes labelled with FAM for Influenza virus A and HEX for Influenza virus B were used at a final concentration of 250 nM. The reaction mix consisted of 2.6 µl of the primer/probe mix, 5 µl ddPCR supermix, 1 µl DTT (300 mM), 2 µl reverse transcriptase, 0.4 µl water, and 9 µl of sample, for a total reaction volume of 20 µl. The RT-ddPCR cycling conditions were as follows: reverse transcription was performed at 50°C for 60 minutes, reverse transcriptase deactivation at 95°C for 10 minutes and denaturation at 94°C for 30 seconds followed by an annealing/extension at 55°C for 1 minute (40 cycles). The final steps included enzyme deactivation at 98°C for 10 minutes and droplet stabilization at 4°C for 30 minutes, followed by an indefinite hold at 4°C. Droplet generation and detection were carried out using the Bio-Rad AutoDG Droplet Digital PCR System, and the thresholds between positive and negative droplets were set with QuantaSoft Software and confirmed manually.

RT-ddPCR for the fecal indicator PMMoV was performed using the following conditions: The final concentrations of forward and reverse primers were 500 nM. The PMMoV probe had a final concentration of 250 nM and was FAM labelled. The reaction mix consisted of 1.2 µl of the primer/probe mix, 5 µl ddPCR supermix, 1 µl DTT (300 mM), 2 µl reverse transcriptase, 1.8 µl water, and 9 µl of sample, for a total reaction volume of 20 µl. The RNA sample was diluted 1:1000 for PMMoV detection. The same cycling conditions as described for IAV and IBV was used.

Data on the compliance with MIQE criteria for ddPCR analyses of IAV, IBV, and PMMoV are provided in the Supplementary Information (Link SI Table 1).

### 2.3 Data Analysis and Statistics

In this study, the Friedman test was chosen as a non-parametric alternative to the repeated measures ANOVA, as it is suitable for comparing multiple groups when the assumption of normality is violated. In this study, the data did not meet the assumptions of normal distribution, which made the Friedman test an appropriate choice (Sheskin, 2003; Hollander et al., 2014). This test ranks the data and assesses differences across related samples, making it particularly suitable for comparing the six investigated methods. Given that RT-ddPCR measurements did not follow a normal distribution, the Friedman test was the appropriate choice for this analysis. Following the Friedman test, the Nemenyi post-hoc test was applied to perform multiple comparisons between the groups. The Nemenyi test is a non-parametric test designed to determine which specific groups differ from each other after finding a significant result in the Friedman test (Nemenyi, Peter Bjorn, 1963; Hollander et al., 2014). It adjusts for multiple comparisons, reducing the risk of Type I errors, and is particularly useful for data that violate parametric test assumptions. This combination of tests allowed for a robust and reliable analysis of the method comparison.

All statistical tests were performed using R version 4.3.1, and p-values below 0.05 were considered significant (*p < 0.05; **p < 0.01; ***p < 0.001). Following packages were used: readxl, dplyr, tidyr, flextable, officer, ggplot2, openxlsx, tidyverse, lubridate, plotrix, ggbreak, gridExtra, svglite.

### 2.4 Virus Spike and Recovery Experiment

The spike-and-recovery experiment was conducted to further validate the best-performing method based on the initial method comparison results. Thus, the method combination PYC/EX1 (PureYield™ Filtration with Maxwell® RSC Instrument) was selected for the spike experiment. In this experiment, a three liters wastewater sample from May 2024 was homogenized for 30 min and then divided into aliquots (4 x 600ml). Inactivated whole IAV and IBV (Zeptometrix, Buffalo, New York, USA) were subsequently spiked into three of the four wastewater samples using three different concentration levels: 10, 10, and 10 gene copies per liter (gc/L). Each aliquot was spiked with both, IAV and IBV, using similar concentrations. The forth 600 ml aliquot was used as a negative control. After thorough mixing, 12x 50-ml aliquots were taken from each sample and used as technical replicates.

This spike experiment was designed to assess the reliability of the PYC/EX1 method and to determine its recovery rate, as it demonstrated the highest efficiency among all tested combinations in the comparative analysis.

## 3 Results

### 3.1 Method Comparison Across Selected Time Points

To evaluate the sensitivity and reliability of different wastewater concentration and extraction methods, six method combinations were tested at four selected time points during the winter season 2023/2024. Each method was assessed for its ability to detect IAV, IBV and PMMoV in wastewater samples collected from four wastewater treatment plants (Fig. 1). The method combination consisting of PureYield™ filtration and Maxwell® RSC extraction (PYC/EX1) displayed the most successful detection rates with respect to the three viruses among all samples representing four distinct WWTPs.

**Figure 1:**
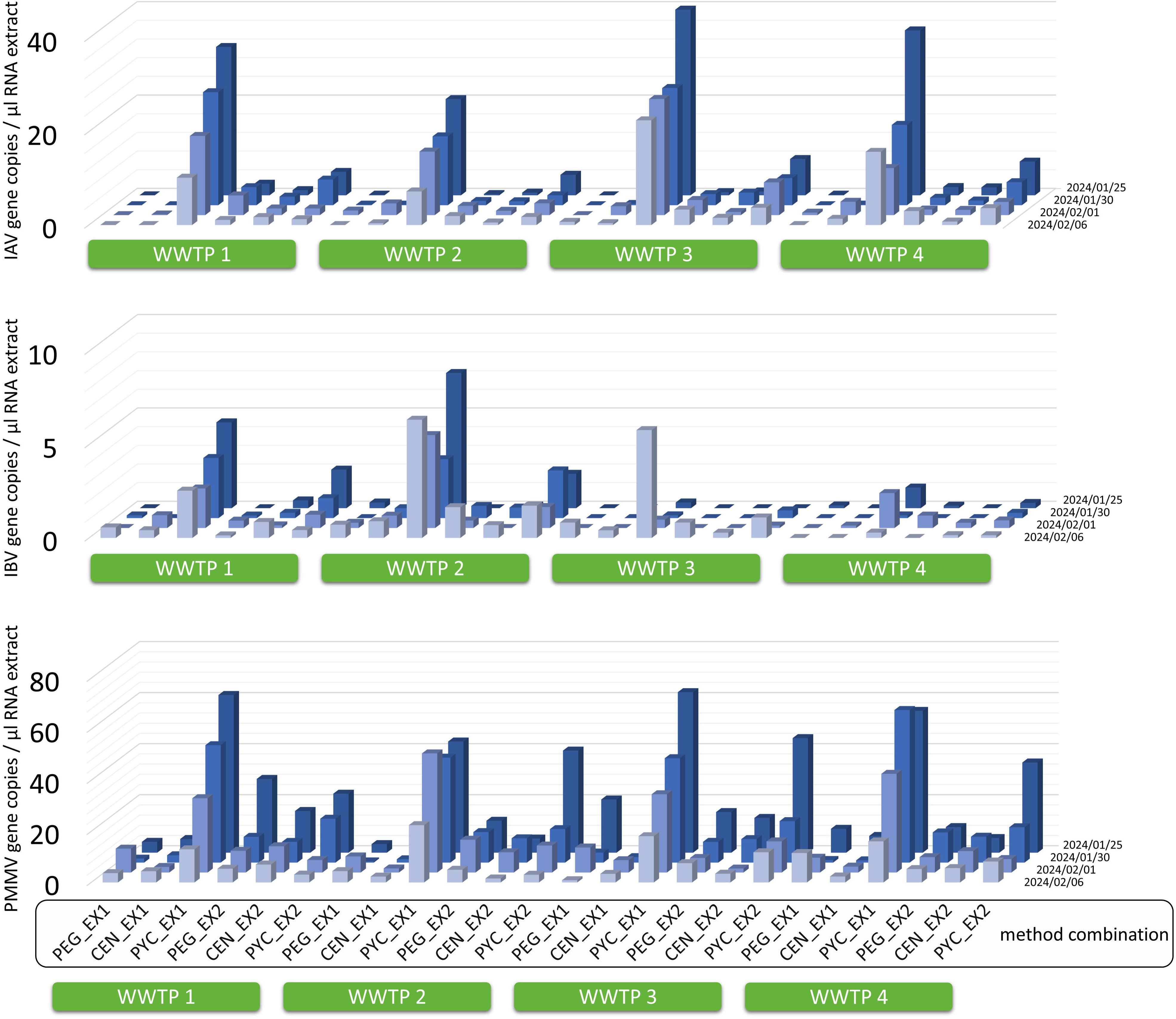
Overview of wastewater sample analyses for Influenza A virus (IAV), Influenza B virus (IBV) and Pepper Mild Mottle Virus (PMMoV) RNA concentrations. Shown are virus gene copies after using six different wastewater concentration and RNA extraction method combinations. Wastewater was sampled at four different wastewater treatment plants (WWTP) and four different time points during the influenza season in winter 2023/2024. PEG: Polyethylene Glycol (PEG) 8000 precipitation, CEN: Centricon™ Plus-70 Filtration, PYC: PureYield™ Filtration, EX1: RNA extraction using the Maxwell® RSC Instrument, EX2: RNA extraction using the InnuPure® C16 touch instrument.

A more detailed analysis of this comparative method assessment is presents in Figs 2-4). As shown in Fig.2 for IAV, the concentration/extraction combinations PYC/EX1, PYC/EX2, and CEN/EX2 yielded positive results for all 16 tested samples (16/16). In contrast, PEG/EX1, CEN/EX1, and PEG/EX2 detected IAV RNA in 3/16, 10/16, and 15/16 samples, respectively (Fig. 2A). The efficiency of the methods was further evaluated by calculating IAV gc/L of wastewater (Fig. 2A, B). The concentration methods PEG, CEN, and PYC combined with the extraction method EX2 detected IAV RNA with mean concentrations ranging from 14,356 to 21,371 gc/L, lacking significant differences across the sample set (Fig. 2B). However, the combination PYC/EX1 achieved the highest mean concentration (24,872 gc/L), significantly exceeding mean values obtained by the other tested combinations (p < 0.001, Fig. 2C). There was no statistically significant difference between EX1 and EX2 when used in combination with PYC (Fig. 2C), highlighting the robustness of both extraction methods with this concentration technique.

**Figure 2:**
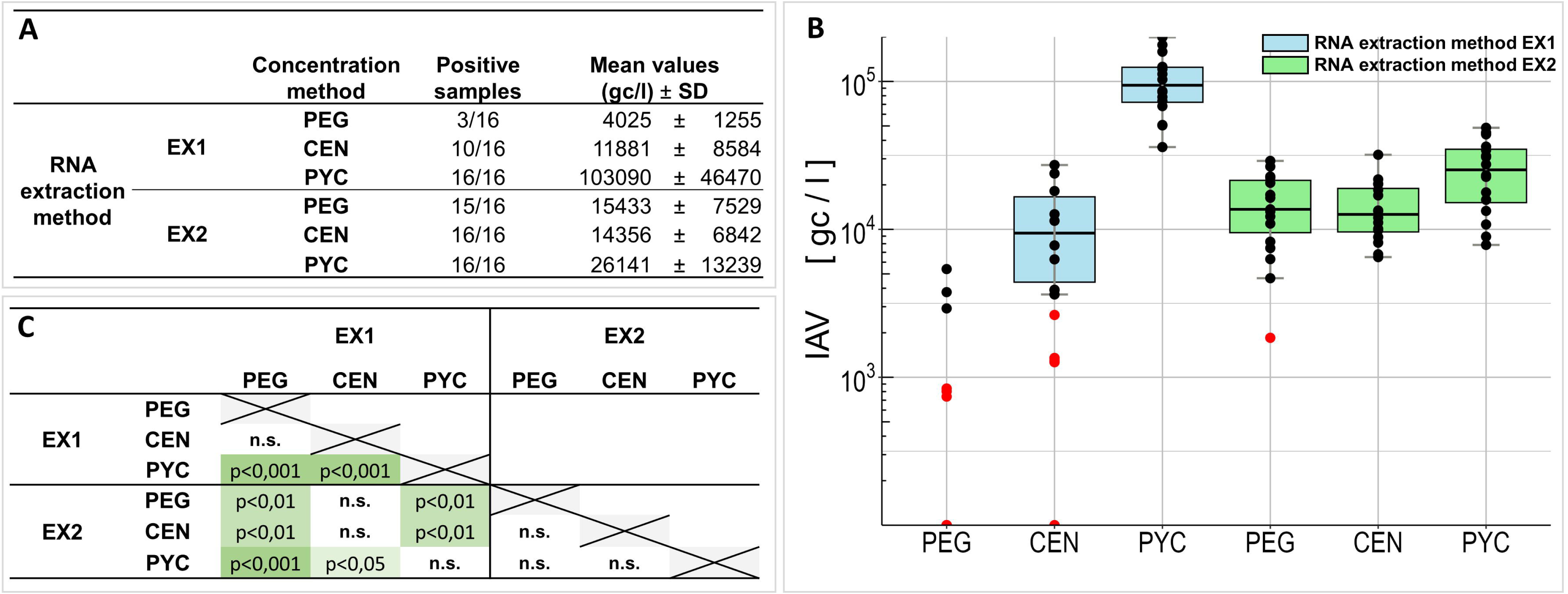
RT-ddPCR based comparison of Influenza A Virus (IAV) component concentration and RNA extraction from wastewater. A and B: Number of IAV positive replicates and mean IAV gene copies and standard deviation (SD) for each method combination. Red dots are indicating measurements below limit of detection (LOQ). C: Results of Nemenyi Post-hoc Test for each method combination; n.s. = not significant, p<0,05 = significant, p<0,01 strong significant, p<0,0001 = very strong significant

For IBV, the concentration/extraction combinations PYC/EX1 and PYC/EX2 yielded positive results in 12/16 and 11/16 samples, respectively (Fig. 3A). Other combinations, such as PEG/EX1, CEN/EX1, and PEG/EX2, achieved fewer positive results, with only 3 to 6 positive samples depending on the combination (Fig. 3A). When efficiency was assessed by calculating IBV gc/L of wastewater, the concentration methods PEG, CEN, and PYC combined with EX2 detected IBV RNA with mean concentrations ranging from 5,252 to 7,631 gc/L (Fig. 3B). The highest mean concentration (16,270 gc/L), achieved by the combination PYC/EX1, was significantly above those achieved by other combinations, except for PYC/EX2 (p < 0.01, Fig. 3C). A general fluctuation in IBV gc across time points was observed, with the lowest or undetectable values associated with the concentration methods PEG and CEN, further emphasizing the reliability of PYC/EX1 and PYC/EX2 for IBV detection.

**Figure 3:**
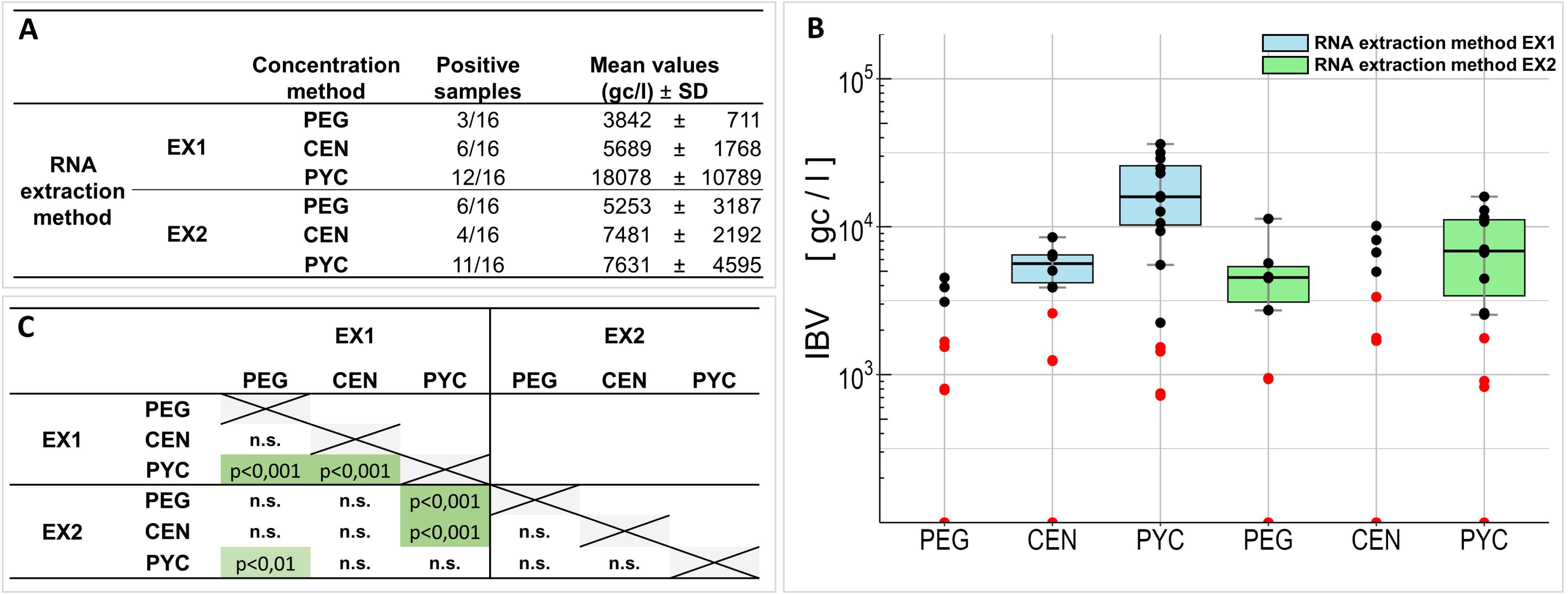
RT-ddPCR based comparison of Influenza B Virus (IBV) component concentration and RNA extraction from wastewater. A and B: Number of IBV positive replicates and mean IBV gene copies and standard deviation (SD) for each method combination. Red dots are indicating measurements below limit of detection (LOQ). C: Results of Nemenyi Post-hoc Test for each method combination; n.s. = not significant, p<0,05 = significant, p<0,01 strong significant, p<0,0001 = very strong significant

PMMoV was measured as a surrogate virus to serve as a fecal control parameter and to evaluate the functionality and efficiency of the tested combinations. Overall, PMMoV was detected in all 16 wastewater samples across all method combinations (Fig. 4). This consistent detection confirms the robustness of the methods and validates their comparability. The mean PMMoV concentrations ranged from 2.6 x 10^7^ gc/L for CEN/EX1 to 1.8 x 10^8^ gc/L for PYC/EX1. Notably, the combination PYC/EX1 yielded the highest average concentrations, with significantly less scatter compared to values achieved by PEG/EX1. The variability observed with PEG/EX1 suggests less consistent recovery efficiency for PMMoV.

**Figure 4:**
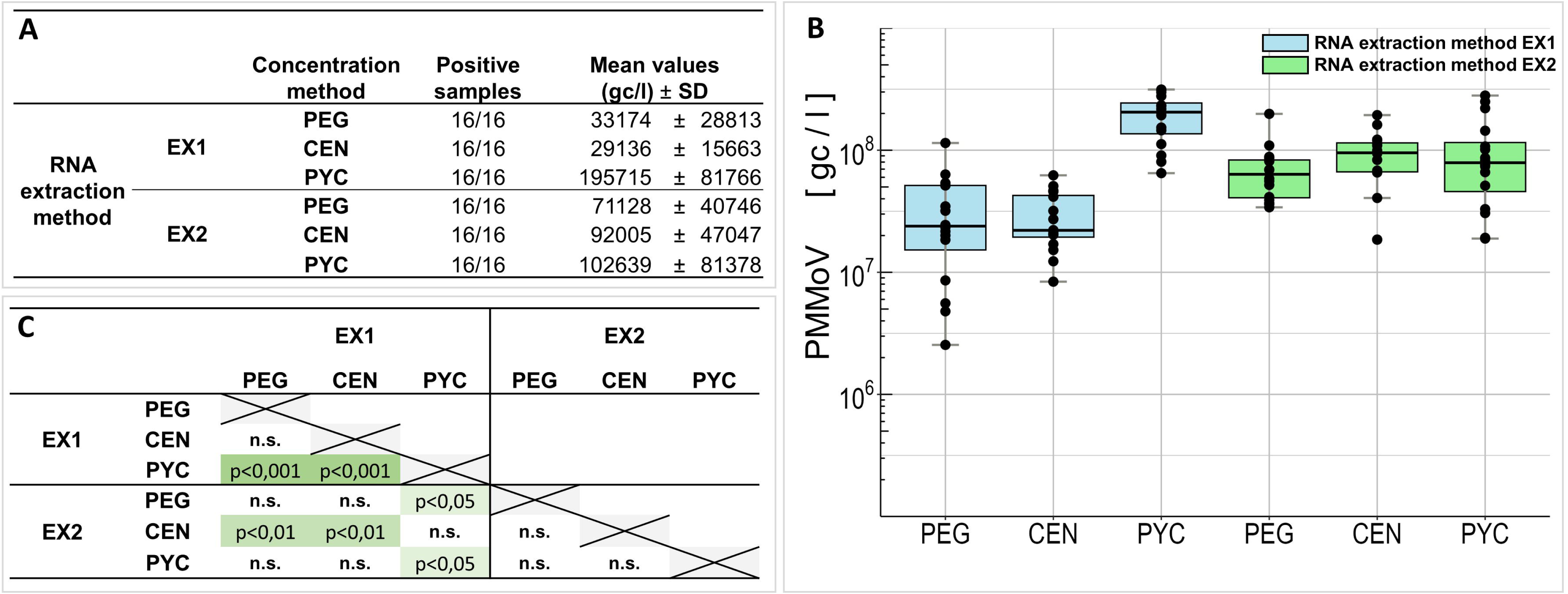
RT-ddPCR based comparison of Pepper Mild Mottle Virus (PMMoV) component concentration and RNA extraction from wastewater. A and B: Number of PMMoV positive replicates and mean PMMoV gene copies and standard deviation (SD) for each method combination. Red dots are indicating measurements below limit of detection (LOQ). C: Results of Nemenyi Post-hoc Test for each method combination; n.s. = not significant, p<0,05 = significant, p<0,01 strong significant, p<0,0001 = very strong significant

Statistical analysis with the Nemenyi post-hoc test confirmed significant differences between PYC/EX1 and each of the other methods (p < 0.05), underscoring the advantage of using optimized protocols for detecting low-abundance viral fragments. This comparison provided a robust foundation for selecting PYC/EX1 as the primary method for subsequent analyses. These results underscore the importance of robust methods like PYC/EX1, which not only demonstrated superior performance for IAV and IBV detection but also ensured stable and reliable PMMoV measurements across all samples. The consistent detection of PMMoV across all combinations reinforces its utility as a fecal control in wastewater-based epidemiology.

### 3.2 Spike Experiment to determine the Recovery Efficiency of Influenza A and B virus

To confirm the reliability of the best-performing method, PYC/EX1, a spike-and-recovery experiment was conducted for both IAV and IBV. In this experiment, inactivated IAV and IBV were spiked into raw wastewater at three concentrations, i.e. 10^4^, 10^5^, and 10^6^ gc/L. Each spiked concentration was tested with 12 replicates, and negative controls confirmed no detectable signal. Results are summarized in Table 2 and Figure 5. For IAV, the recovery rates varied by spike level, with an average recovery of: 80.4% at 10^4^ gc/L, 69.7% at 10^5^ gc/L, and 63.4% at 10^6^ gc/L. The coefficient of variation (CV) ranged from 41.4% at the lowest concentration to 32.8% at the highest concentration, indicating consistent recovery performance across spike levels (Table 2; Fig. 5A). For IBV, the recovery rates were slightly lower exhibiting 72.3% at 10^4^ gc/L, 64.9% at 10^5^ gc/L, and 60.3% at 10^6^ gc/L. The coefficient of variation ranged from 28.0% at 10^4^ gc/L to 36.3% at 10^6^ gc/L, showing a similar trend of decreasing recovery rates with increasing spike concentrations (Table 2; Fig.5B). The practical limit of quantification (LOQ) for PYC/EX1 was established at 2,261 gc/L, confirming the method’s capability for reliable detection and quantification in complex wastewater matrices. These results validate the effectiveness of PYC/EX1 for the quantitative detection of both IAV and IBV in wastewater. The methodical workflow demonstrated high recovery rates, low variability, and robustness across different spike levels. Detailed raw data for this experiment are documented in the Supplementary Information (LINK SI Table 3).

**Figure 5:**
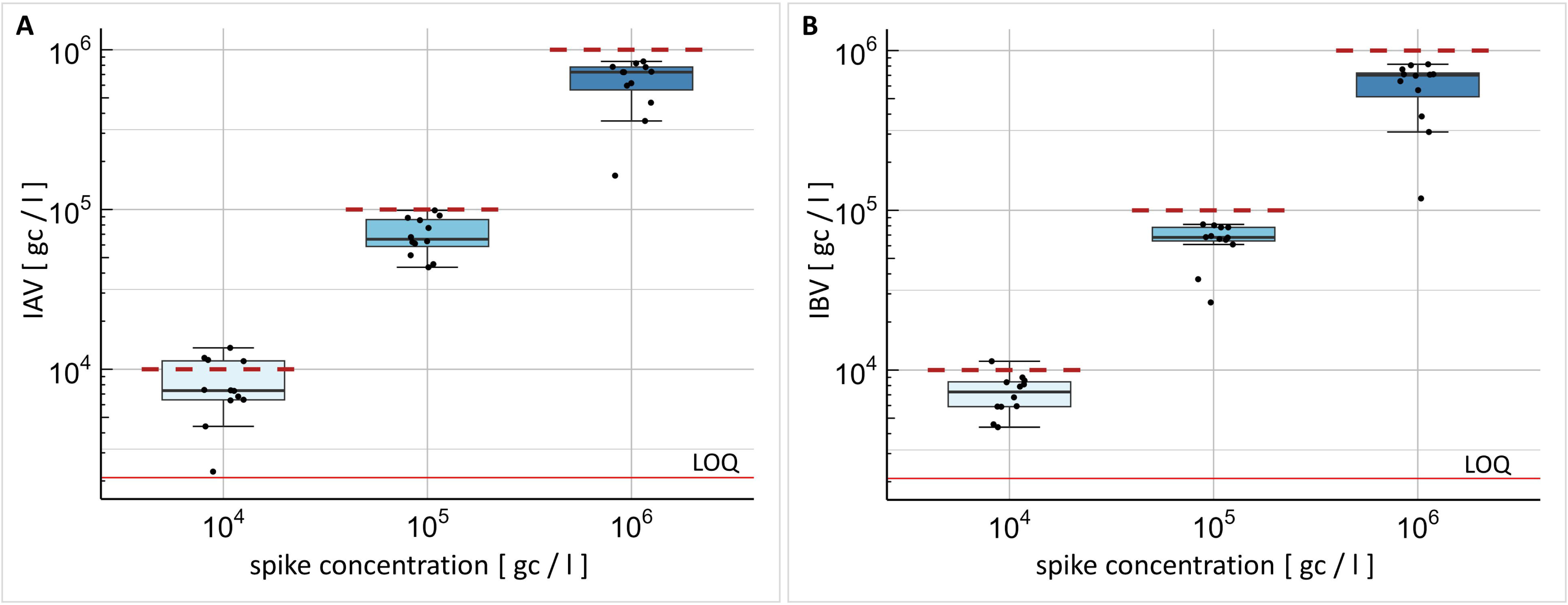
Boxplots representing Influenza virus gene copies from spiked wastewater. Method combination PYC/EX1 (PureYield™ Filtration/Maxwell® RSC Instrument RNA extraction) was used to extract 10^4^ to 10^6^ spike-in gene copies per liter (red dashed line; each concentration in triplicates). LOQ: Limit of quantification. Raw data can be found in Supplementary information.

**Table 2:** Mean values, recovery rate and variation coefficient for Influenza A and B viruses. abbreviations: gc = gene copies, IAV = influenza A virus, IBA = influenza B virus.

|  | spike concentration<br>[gc/l] | Mean values<br>[gc/l] | recovery rate<br>[%] | variation coefficient<br>[%] |
| --- | --- | --- | --- | --- |
| IAV | 10<sup>4</sup> | 8,0 x 10<sup>3</sup> | 80,4 | 41,4 |
|  | 10<sup>5</sup> | 6,9 x 10<sup>4</sup> | 69,7 | 26,6 |
|  | 10<sup>6</sup> | 6,3 x 10<sup>5</sup> | 63,4 | 32,8 |
| IBV | 10<sup>4</sup> | 7,2 x 10<sup>3</sup> | 72,3 | 28,0 |
| | $10^5$ | $6,5 \times 10^4$ | 64,9 | 26,2 |
| | $10^6$ | $6,0 \times 10^5$ | 60,3 | 36,3 |

### 3.3 Viral Load Trends for IAV, IBV, and PMMoV Across the Winter Season

Using the validated PYC/EX1 method, we monitored viral loads for IAV, IBV, and PMMoV in wastewater throughout the winter season 2023 / 2024 across four WWTPs located in Mecklenburg Western Pomerania, Germany. Seasonal trends for IAV and IBV in each WWTP are presented in Figure 6 and data for PMMoV are listed in the Supplementary Information (LINK SI Table 4). Periodic increases in viral loads were observed, corresponding to known Influenza peaks during the winter season. These increases were consistent across all WWTPs, with some variation in magnitude, likely reflecting differences in local population size and Influenza activity. PMMoV was consistently detected at high concentrations throughout the monitoring period. The stable presence of PMMoV across all samples confirmed sample integrity and validated the comparability of the data collected across the four WWTPs. For all IAV/IBV analyses, raw data points were included in the graphical representations to provide transparency in the observed values. Loess regression curves with a span of 0.7 were applied to highlight trends over time, balancing smoothing with data fidelity. This approach allowed for reliable and continuous viral monitoring while accounting for day-to-day variability in wastewater viral loads.

**Figure 6:**
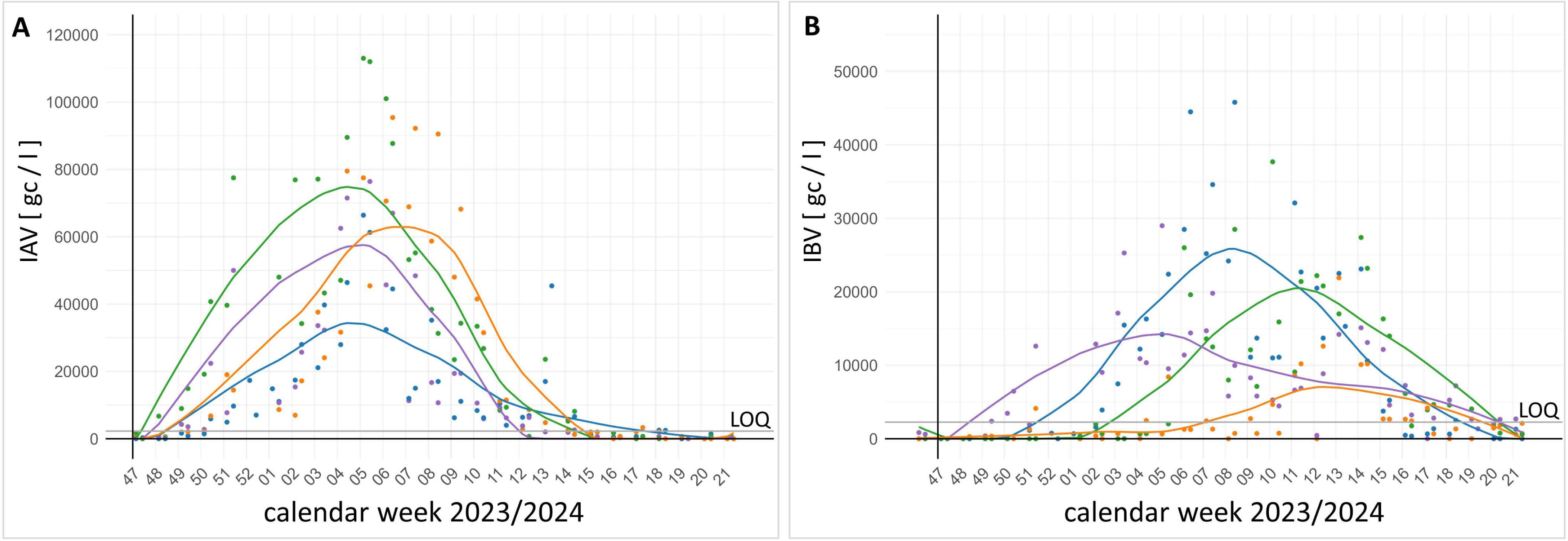
Influenza virus gene copies detected from four wastewater treatment plants in Mecklenburg Western Pomerania, Germany during the influenza season 2023/2024. Each wastewater treatment plant has one color (blue, orange, green or violet). The dots are indicating the calculated gene copies per liter and the lines are calculated by using the LOESS regression. Limit of detection (LOQ) is shown as a grey line. A. Influenza A virus (IAV) gene copies per liter winter season 2023/2024. B. Influenza B virus (IAV) gene copies per liter winter season 2023/2024.

### 3.4 Simulation Using Factor-Based Adjustment

To illustrate the difference in detection sensitivity between PYC/EX1 and the other method combinations, a scaling factor was calculated. This factor represents the average difference in detection levels achieved by PYC/EX1 compared to the other methods over the study period. Using this adjustment, viral loads were recalculated to simulate detection outcomes if the other methods had been employed instead of PYC/EX1. The simulation provides a comparative prediction of how detection timing and sensitivity would differ across the tested methods. The simulated aggregated detection levels for IAV across the four tested WWTPs in in the study region are shown in Figure 7A1. The blue curve represents actual IAV concentrations measured with PYC/EX1, smoothed using Loess regression. Dotted lines indicate the recalculated concentrations for the other methods (PYC/EX2, PEG/EX2, CEN/EX2, CEN/EX1, and PEG/EX1) based on their mean differences from PYC/EX1. The grey area highlights the range of concentrations expected if less sensitive methods had been used, emphasizing the variability in detection levels. A key observation is the delayed breakthrough of the limit of quantification (LOQ) for the other methods compared to PYC/EX1 (Fig. 7A1). The magnified view of Figure 7A2 illustrates the early rise of the IAV presence and the differences in detection timing. The use of PYC/EX1 allowed for detection approximately one week earlier than the second-best method (PYC/EX2). This observation is important to critically interpret wastewater signals when using them for early-warning surveillance systems. Similar trends were observed for IBV, as shown in Figure 7B1 and 7B2. The blue line in this figure represents IBV RNA concentrations measured with PYC/EX1, while the dotted lines depict recalculated concentrations for the other methods. The grey-shaded area highlights the range of concentrations if alternative methods had been employed. The grey horizontal line represents the LOQ for PYC/EX1, providing a reference point for method sensitivity. As with IAV, the simulation demonstrates a significant lead time for detection using PYC/EX1. This earlier detection window is particularly important for tracking IBV during seasonal peaks, allowing for identification of rising virus loads up to two weeks earlier compared to less sensitive methods.

**Figure 7:**
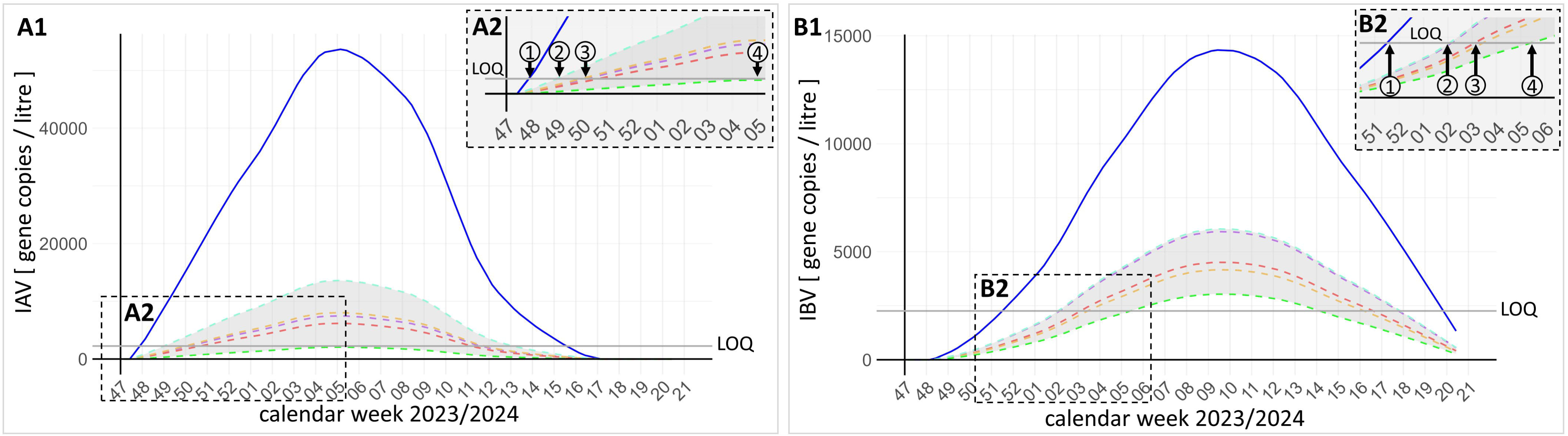
Scaling factor-based simulation of method combination performance compared to the most sensitive combination PYC_EX1 (PureYield™ Filtration / Maxwell® RSC Instrument RNA extraction) to illustrate the time frame of early virus detection. Shown are Influenza A virus (IAV) (Fig.7A1) and Influenza B Virus (IBV) (Fig.7B1) gene copies for aggregated four investigated wastewater treatment plants located in Mecklenburg Western Pomerania, Germany. The blue line represents LOESS regression of Influenza virus gene copies of all four wastewater treatment plants. The dashed lines in light blue, violet, red, orange and green are representing the calculated values for the other tested methods in this study. The grey area indicates the range between the second best and worst method combination. Limit of detection (LOQ) is shown as a grey line. Figures 7A2 and 7B2 show the different times at which the different method combinations for IAV and IBV would be above the LOQ.

## 4 Discussion

Precise and accurate quantification of viral nucleic acid copies in wastewater is essential for the successful application of WBE. Overall, this study highlights the importance of comparing multiple methods to ensure reliable and reproducible results. By systematically evaluating six method combinations, we demonstrated how differences in sensitivity and efficiency can impact the detection and quantification of viral targets, including IAV, IBV and PMMoV. Comparison of methods is particularly critical for low-abundance targets like IBV, where small variations in sensitivity could lead to false-negative results. Conversely, robust methods are necessary to avoid overestimations and false-positive interpretations for high-abundance targets like PMMoV. This underscores the need for method validation tailored to specific WBE goals, such as early detection of emerging pathogens or tracking seasonal trends of endemic viruses. Ultimately, the careful selection and comparison of methods help to minimize uncertainties and to ensure that WBE systems can reliably detect viral signals at an early stage, enabling timely public health interventions.

### 4.1 Method Combination Performance

Comparison of methods for virus detection in wastewater has been a key focus in WBE research. Numerous studies have evaluated individual methods for virus concentration and extraction, particularly in the context of emerging pathogens like SARS-CoV-2 (Dumke et al., 2021; Forés et al., 2021; Pearson et al., 2021; Othman et al., 2023; Antkiewicz et al., 2024). However, limited data exist on the performance of method combinations, which integrate both concentration and extraction steps, and their impact on the detection of specific viral targets. The inclusion of both IAV and IBV as targets addresses a critical gap in the existing literature, as most prior studies have focused on high-abundance surrogate markers or emerging pathogens with higher stability (Farkas et al., 2020; Boehm et al., 2023b). Building on previous studies that evaluated individual methods, our study highlights that comparing method combinations is crucial to understanding how different protocols influence detection sensitivity, recovery efficiency, and overall reliability, particularly for low-abundance targets like e.g. IBV. By investigating six different method combinations, this study aims to identify protocols that provide robust and consistent results, minimize false negatives, and ensure accurate viral quantification. We chose these combinations of methods because they are already used in the nationwide wastewater surveillance for SARS-CoV-2 in Germany (Dumke et al., 2022; Marquar et al., 2024). The evaluation of such methods is important to ensure the reliability of the detection methods and to minimize inaccuracies in terms of precision and accuracy. Furthermore, this comparison is especially important for developing early-warning systems that rely on precise and timely detection of viral signals in wastewater, enabling public health measures such as targeted vaccination campaigns to be implemented within the calculated time frame.

The results across all three targets - IAV, IBV, and PMMoV - consistently show that PYC-based methods outperform both PEG and CEN combinations tested in this study. This trend holds true across both extraction protocols (EX1 and EX2), but the most striking performance was observed with PYC_EX1, which consistently yielded the highest recovery rates and the most reliable results for all targets. The superior performance of PYC methods may reflect their ability to efficiently concentrate viral particles while minimizing the loss of RNA during the concentration step. This is particularly critical for low-abundance targets like IBV, where even minor losses can result in concentrations near or below the detection limit. The consistent results for PMMoV across all methods highlight its robustness as a process control but also underscore the relative challenges associated with influenza virus detection. In contrast, PEG and CEN methods showed reduced recovery rates, particularly for IBV. This finding contrasts with previous studies where PEG precipitation has been highlighted as a robust method for viral concentration (Dumke et al., 2022; Farkas et al., 2022). The reduced performance observed here could stem from target-specific challenges, such as the fragility of influenza viruses or matrix-specific inhibitors that affect PEG efficiency in these samples. While no significant differences between EX1 and EX2 were observed for IAV and IBV, the findings for PMMoV indicate that EX1 can outperform EX2 in certain scenarios. The higher recovery rates observed with PYC/EX1 compared to PYC/EX2 highlight the potential impact of subtle differences in extraction protocols, such as lysis efficiency, inhibitor removal, or elution conditions as discussed before (Ahmed et al., 2021; Barril et al., 2021; Gouthro et al., 2024; Linzner et al., 2024).

Across all targets, PYC/EX1 provided the highest sensitivity and recovery efficiency, making it the optimal choice for WBE applications. The significant advantage of PYC/EX1 over PYC/EX2 for PMMoV also highlights its broader utility for robust virus detection. The observed difference between PYC/EX1 and PYC/EX2 for PMMoV suggests that even small adjustments in extraction protocols can affect recovery efficiency. Laboratories should validate extraction protocols not only for target sensitivity but also for consistency across multiple viral markers. The reduced performance of PEG and CEN methods reinforces the need for optimization, particularly for low-abundance or fragile targets. While these methods remain viable for high-abundance targets like PMMoV, their limitations should be carefully considered in WBE workflows.

### 4.2 Spike Experiment and Sensitivity Validation

The spike-and-recovery experiment serves as a critical validation tool for assessing the reliability of methods used in WBE. Recovery rates observed in this study for IAV and IBV ranged between 60.3% and 80.4%, with coefficients of variation (CV) from 28.0% to 41.4%. These values are consistent with or exceed recovery rates reported in prior studies for other RNA viruses in wastewater, such as SARS-CoV-2, which have shown recovery efficiencies typically ranging from 4% to approximately 100% depending on the method and matrix (Dumke et al., 2021; Fonseca et al., 2022; Zheng et al., 2022; Toribio-Avedillo et al., 2023, 2024). The relatively high recovery rates observed here underscore the robustness of the evaluated protocol for detecting low-abundance targets like influenza viruses. However, variability in recovery rates across spike levels highlights the influence of sample matrix and methodological details, such as the efficiency of lysis and RNA extraction steps. While these recovery rates are suitable for quantitative applications, they also point to the need for caution in interpreting absolute viral concentrations. Differences in recovery efficiency can lead to under- or overestimations of viral loads, which must be accounted for when drawing conclusions about viral prevalence or trends. The practical implications of these findings for WBE applications are significant. Reliable recovery ensures that even low concentrations of viral RNA, such as those observed for IBV in this study, can be detected and quantified. Ensuring sensitivity at low viral loads is a key aspect of wastewater-based early-warning systems, as it enables more timely and effective public health responses. However, the variability observed also reinforces the importance of integrating recovery controls and standardized protocols into routine surveillance workflows to minimize uncertainties. From a comparative perspective, these results contribute to a growing body of evidence emphasizing the importance of method validation for each target virus. While surrogate viruses like PMMoV offer valuable process controls, their recovery efficiencies do not necessarily predict the performance of methods for other targets. The distinct challenges posed by influenza viruses, including their lower abundance and RNA stability in wastewater, highlight the need for tailored validation studies to ensure that methods are fit for purpose. In conclusion, the spike-and-recovery experiment demonstrates the feasibility of reliably detecting IAV and IBV in wastewater. Nevertheless, it also underscores the need for continuous refinement of methods and cautious interpretation of quantitative data, particularly when extrapolating trends or making public health decisions based on viral load estimates.

### 4.3 Public Health Implications

The findings of this study underscore the critical need to select and validate method combinations for WBE, especially when targeting temporarily low-frequency viruses like IAV and IBV. PYC/EX1 emerged as the most reliable and sensitive method across all tested targets, but the observed variability among other combinations highlights the necessity of tailoring methods to specific surveillance objectives. Such findings emphasize the critical role of early detection in public health planning and intervention. Additionally, timely detection enables public health advisories to be issued to healthcare providers and communities, raising awareness and promoting preventative measures (Gahlot et al., 2023). Furthermore, early warning systems facilitate the prioritization of medical and logistical resources, ensuring that regions showing signs of increasing viral loads are adequately prepared to manage potential outbreaks (Clark et al., 2023; Grassly et al., 2025). The observed variability in performance among the tested method combinations highlights the need for further research to improve the reliability and applicability of WBE. First, traditional concentration methods such as PEG precipitation and Centricon filtration require optimization to enhance their efficiency for low-abundance targets like IAV and IBV. This could involve integrating additional steps, such as enzymatic lysis or improved buffer formulations, to increase RNA recovery rates and reduce variability. Second, high-performing methods like PYC/EX1, which demonstrated consistent sensitivity and reliability in this study, should be validated across diverse wastewater matrices and environmental conditions. Such validation would ensure that these methods perform robustly in different geographical and operational contexts, supporting their broader implementation in WBE programs. Finally, the development of standardized protocols is essential to improve the comparability of WBE data across regions and studies. Standardization would facilitate more accurate meta-analyses, enhance data integration, and provide a clearer picture of viral trends at local, national, and global levels. By addressing these challenges, WBE can evolve into an even more reliable and impactful tool for public health surveillance.

## Data Availability

All data produced in the present work are contained in the manuscript

## 5 Conflict of Interest

The authors declare that the research was conducted in the absence of any commercial or financial relationships that could be construed as a potential conflict of interest.

## 6 Author Contributions

TF: Conceptualization, Data curation, Formal analysis, Investigation, Methodology, Validation, Visualization, Writing – original draft. EEZ: Data curation, Formal analysis, Investigation, Methodology. BS: Methodology. ML: Project administration, Writing – review & editing. UB: Funding acquisition, Project administration, Writing – review & editing. TG: Funding acquisition, Project administration, JS: Funding acquisition, Project administration, HCS: Conceptualization, Methodology, Writing – review & editing. SS: Formal analysis, Investigation, Methodology. AA: Formal analysis, Investigation, Methodology. BW: Conceptualization, Funding acquisition, Project administration, Supervision, Validation, Visualization, Writing – original draft. RK: Conceptualization, Supervision, Validation, Visualization, Writing – original draft.

## 7 Funding

This work was funded and supported by the German Federal Ministry of Health as part of the project “Abwassermonitoring für die epidemiologische Lagebewertung” (AMELAG).

## 8 Acknowledgments

We sincerely appreciate the support of the wastewater treatment plant operators and staff for providing the wastewater samples, with special thanks to the wastewater disposal companies in Mecklenburg-Vorpommern - Nordwasser GmbH, Schweriner Abwasserentsorgung Eigenbetrieb der Landeshauptstadt Schwerin, Neubrandenburger Wasserbetriebe GmbH, and Universitäts-und Hansestadt Greifswald and Stadtwerke Greifswald GmbH (Abwasserwerk). In addition, we would especially like to thank our RKI colleagues Udo Buchholz and Susan Abunjiela for their useful comments and advice and our UBA colleagues Cristina J. Saravia and Alexander Kerndorff for their help with providing the samples for the recovery rate experiments. We would like to thank the entire AMELAG team for the fruitful discussions and helpful input.

## 10 Data Availability Statement

The original contributions presented in the study are included in the article/Supplementary Material, further inquiries can be directed to the corresponding authors

## 11 Supplementary Material

## Notes

### Competing Interest Statement

The authors have declared no competing interest.

### Funding Statement

This work was funded and supported by the German Federal Ministry of Health as part of the project - Abwassermonitoring fuer die epidemiologische Lagebewertung (AMELAG).

