## Supplementary_Information for "Comparative assessment of combined concentration and extraction methods for Influenza A and B virus detection in wastewater"

Table 1. Additional Information for ddPCR according to MIQE-Guidlines

| Primer/Probe target, positive controls | Description | Sequence (5’-3’) |
| --- | --- | --- |
| Influenza A virus | IAV For1 | CAA GAC CAA TCY TGT CAC CTC TGA C |
|  | IAV Rev1 | GCA TTY TGG ACA AAV CGT CTA CG |
|  | IAV Probe | FAM-TGC AGT CCT CGC TCA CTG GGC ACG-BHQ-1 |
| Influenza B virus | IBV For1 | TCC TCA AYT CAC TCT TCG AGC G |
|  | IBV Rev1 | CGG TGC TCT TGA CCA AAT TGG |
|  | IBV Probe | HEX-CCA ATT CGA GCA GCT GAA ACT GCG GTG-BHQ-1 |
| Positive control for Influenza A virus | IAV Pos | GGAGCAGACAAGCCCGTCAGGGCGCGTCAGCGGGTGTTGGCGGGTGTCGGGGCTGGCTTAACTATGCGGCATCAGAGCAGATTGTACTGAGAGAAAGGCAATTGGGTACCGAGCTCGCGGCCGCAAGCAAAGACAAGACCAATCCTGTCACCTCTGACTAAGGGGATTTTAGGGTTTGTGTTCACGCTCACCGTGCCCAGTGAGCGAGGACTGCAGCGTAGACGCTTTGTCCAAAATGCCCTAAACCTGCTTTTGCTCGCTTGGATCCGAATTCAAAGGTGAAATTGTTATCCGCTCACAATTCCACACAACATACGAGCCGGAAGCATAAAGTGTAAAGCCTG |
| Positive control for Influenza B virus | IBV Pos | GGAGCAGACAAGCCCGTCAGGGCGCGTCAGCGGGTGTTGGCGGGTGTCGGGGCTGGCTTAACTATGCGGCATCAGAGCAGATTGTACTGAGAGAAAGGCAATTGGGTACCGAGCTCGCGGCCGCAAGCTCGGATCCTCAATTCACTCTTCGAGCGTCTTAATGAAGGACATTCAAAGCCAATTCGAGCAGCTGAAACTGCGGTGGGAGTCTTATCCCAATTTGGTCAAGAGCACCGACTATACCTGCTTTTGCTCGCTTGGATCCGAATTCAAAGGTGAAATTGTTATCCGCTCACAATTCCACACAACATACGA  GCCGGAAGCATAAAGTGTAAAGCCTG |
| PMMoV | PMMoV-F | GAG TGG TTT GAC CTT AAC GTT TGA |
|  | PMMoV-R | TTG TCG GTT GCA ATG CAA GT |
|  | PMMoV-Probe | FAM-CCT ACC GAA GCA AAT G-TAMRA |
| Positive control for PMMoV | PMMoV-Pos | GGA TGT GTA ATA CAT TAG GCG TAG ATC CAT TGG TGG CAG CAA AGG TAA TGG TAG CTG TGG TTT CAA ATG AGA GTG GTT TGA CCT TAA CGT TTG AGA GGC CTA CCG AAG CAA ATG TCG CAC TTG CAT TGC AAC CGA CAA TTA CAT CAA AGG AGG AAG GTT CGT TGA AGA TTG TGT CGT CAG ACG TAG GTG AGT CCT CAA TCA AGG AAG |

Table 2. Raw data of method Comparison Across Selected Time Points

| **Date** | **WTTP** | **Method combination** | **PMMoV** | **IAV** | **IBV** |
| --- | --- | --- | --- | --- | --- |
|  |  |  | **[gene copies / µl RNA extract]** | | |
| 25.01.2024 | 1 | PYC/EX1 | 61,78 | 31,78 | 4,60 |
| 30.01.2024 | 1 | PYC/EX1 | 46,00 | 24,22 | 3,22 |
| 01.02.2024 | 1 | PYC/EX1 | 29,11 | 16,98 | 2,12 |
| 06.02.2024 | 1 | PYC/EX1 | 12,98 | 10,16 | 2,53 |
| 25.01.2024 | 1 | PYC/EX2 | 23,11 | 5,02 | 2,07 |
| 30.01.2024 | 1 | PYC/EX2 | 17,18 | 5,49 | 1,07 |
| 01.02.2024 | 1 | PYC/EX2 | 4,89 | 1,43 | 0,71 |
| 06.02.2024 | 1 | PYC/EX2 | 3,07 | 1,26 | 0,42 |
| 25.01.2024 | 1 | PEG/EX1 | 4,24 | 0,00 | 0,00 |
| 30.01.2024 | 1 | PEG/EX1 | 1,56 | 0,00 | 0,14 |
| 01.02.2024 | 1 | PEG/EX1 | 9,36 | 0,00 | 0,00 |
| 06.02.2024 | 1 | PEG/EX1 | 3,64 | 0,00 | 0,56 |
| 25.01.2024 | 1 | PEG/EX2 | 28,89 | 2,49 | 0,00 |
| 30.01.2024 | 1 | PEG/EX2 | 10,09 | 3,87 | 0,14 |
| 01.02.2024 | 1 | PEG/EX2 | 8,51 | 4,22 | 0,40 |
| 06.02.2024 | 1 | PEG/EX2 | 5,38 | 1,09 | 0,14 |
| 25.01.2024 | 1 | CEN/EX1 | 5,36 | 0,66 | 0,13 |
| 30.01.2024 | 1 | CEN/EX1 | 2,87 | 0,00 | 0,00 |
| 01.02.2024 | 1 | CEN/EX1 | 2,19 | 0,14 | 0,68 |
| 06.02.2024 | 1 | CEN/EX1 | 4,38 | 0,00 | 0,41 |
| 25.01.2024 | 1 | CEN/EX2 | 16,33 | 1,12 | 0,42 |
| 30.01.2024 | 1 | CEN/EX2 | 8,11 | 1,83 | 0,28 |
| 01.02.2024 | 1 | CEN/EX2 | 10,29 | 1,43 | 0,14 |
| 06.02.2024 | 1 | CEN/EX2 | 7,02 | 1,71 | 0,85 |
| 25.01.2024 | 2 | PYC/EX1 | 43,56 | 20,62 | 7,24 |
| 30.01.2024 | 2 | PYC/EX1 | 41,11 | 14,76 | 3,16 |
| 01.02.2024 | 2 | PYC/EX1 | 46,67 | 13,62 | 4,98 |
| 06.02.2024 | 2 | PYC/EX1 | 22,44 | 7,20 | 6,33 |
| 25.01.2024 | 2 | PYC/EX2 | 40,00 | 4,40 | 1,84 |
| 30.01.2024 | 2 | PYC/EX2 | 13,11 | 2,13 | 2,56 |
| 01.02.2024 | 2 | PYC/EX2 | 10,58 | 2,53 | 1,12 |
| 06.02.2024 | 2 | PYC/EX2 | 3,02 | 1,73 | 1,73 |
| 25.01.2024 | 2 | PEG/EX1 | 3,36 | 0,15 | 0,30 |
| 30.01.2024 | 2 | PEG/EX1 | 0,46 | 0,13 | 0,00 |
| 01.02.2024 | 2 | PEG/EX1 | 6,31 | 0,98 | 0,28 |
| 06.02.2024 | 2 | PEG/EX1 | 4,44 | 0,00 | 0,71 |
| 25.01.2024 | 2 | PEG/EX2 | 12,51 | 0,27 | 0,00 |
| 30.01.2024 | 2 | PEG/EX2 | 11,96 | 0,92 | 0,65 |
| 01.02.2024 | 2 | PEG/EX2 | 12,84 | 1,99 | 0,40 |
| 06.02.2024 | 2 | PEG/EX2 | 4,96 | 1,92 | 1,65 |
| 25.01.2024 | 2 | CEN/EX1 | 4,91 | 0,41 | 0,27 |
| 30.01.2024 | 2 | CEN/EX1 | 1,30 | 0,13 | 0,53 |
| 01.02.2024 | 2 | CEN/EX1 | 1,60 | 2,51 | 0,66 |
| 06.02.2024 | 2 | CEN/EX1 | 2,29 | 0,38 | 0,89 |
| 25.01.2024 | 2 | CEN/EX2 | 5,53 | 0,57 | 0,14 |
| 30.01.2024 | 2 | CEN/EX2 | 9,51 | 0,85 | 0,56 |
| 01.02.2024 | 2 | CEN/EX2 | 7,93 | 0,94 | 0,00 |
| 06.02.2024 | 2 | CEN/EX2 | 1,56 | 0,55 | 0,68 |
| 25.01.2024 | 3 | PYC/EX1 | 62,89 | 39,78 | 0,31 |
| 30.01.2024 | 3 | PYC/EX1 | 40,89 | 25,11 | 0,15 |
| 01.02.2024 | 3 | PYC/EX1 | 30,67 | 24,89 | 0,45 |
| 06.02.2024 | 3 | PYC/EX1 | 18,11 | 22,44 | 5,78 |
| 25.01.2024 | 3 | PYC/EX2 | 44,89 | 7,78 | 0,00 |
| 30.01.2024 | 3 | PYC/EX2 | 16,24 | 5,80 | 0,41 |
| 01.02.2024 | 3 | PYC/EX2 | 12,20 | 7,02 | 0,13 |
| 06.02.2024 | 3 | PYC/EX2 | 11,82 | 3,71 | 1,10 |
| 25.01.2024 | 3 | PEG/EX1 | 20,84 | 0,00 | 0,00 |
| 30.01.2024 | 3 | PEG/EX1 | 3,93 | 0,00 | 0,00 |
| 01.02.2024 | 3 | PEG/EX1 | 9,80 | 0,00 | 0,00 |
| 06.02.2024 | 3 | PEG/EX1 | 0,87 | 0,68 | 0,82 |
| 25.01.2024 | 3 | PEG/EX2 | 15,91 | 0,68 | 0,00 |
| 30.01.2024 | 3 | PEG/EX2 | 8,13 | 2,38 | 0,00 |
| 01.02.2024 | 3 | PEG/EX2 | 5,69 | 3,24 | 0,00 |
| 06.02.2024 | 3 | PEG/EX2 | 7,56 | 3,31 | 0,82 |
| 25.01.2024 | 3 | CEN/EX1 | 1,79 | 0,82 | 0,00 |
| 30.01.2024 | 3 | CEN/EX1 | 2,14 | 0,28 | 0,00 |
| 01.02.2024 | 3 | CEN/EX1 | 4,82 | 1,91 | 0,00 |
| 06.02.2024 | 3 | CEN/EX1 | 3,36 | 0,41 | 0,41 |
| 25.01.2024 | 3 | CEN/EX2 | 13,62 | 0,83 | 0,00 |
| 30.01.2024 | 3 | CEN/EX2 | 9,20 | 2,69 | 0,00 |
| 01.02.2024 | 3 | CEN/EX2 | 1,56 | 0,68 | 0,00 |
| 06.02.2024 | 3 | CEN/EX2 | 3,42 | 1,56 | 0,28 |
| 25.01.2024 | 4 | PYC/EX1 | 55,56 | 35,33 | 1,10 |
| 30.01.2024 | 4 | PYC/EX1 | 59,78 | 17,22 | 0,14 |
| 01.02.2024 | 4 | PYC/EX1 | 38,67 | 10,09 | 1,87 |
| 06.02.2024 | 4 | PYC/EX1 | 16,09 | 15,69 | 0,29 |
| 25.01.2024 | 4 | PYC/EX2 | 35,33 | 7,22 | 0,28 |
| 30.01.2024 | 4 | PYC/EX2 | 13,84 | 4,93 | 0,28 |
| 01.02.2024 | 4 | PYC/EX2 | 5,27 | 2,84 | 0,41 |
| 06.02.2024 | 4 | PYC/EX2 | 8,20 | 3,62 | 0,15 |
| 25.01.2024 | 4 | PEG/EX1 | 9,36 | 0,15 | 0,15 |
| 30.01.2024 | 4 | PEG/EX1 | 1,01 | 0,00 | 0,00 |
| 01.02.2024 | 4 | PEG/EX1 | 5,80 | 0,53 | 0,00 |
| 06.02.2024 | 4 | PEG/EX1 | 11,51 | 0,00 | 0,00 |
| 25.01.2024 | 4 | PEG/EX2 | 10,02 | 1,78 | 0,14 |
| 30.01.2024 | 4 | PEG/EX2 | 11,82 | 1,59 | 0,00 |
| 01.02.2024 | 4 | PEG/EX2 | 6,02 | 1,20 | 0,67 |
| 06.02.2024 | 4 | PEG/EX2 | 5,24 | 3,00 | 0,00 |
| 25.01.2024 | 4 | CEN/EX1 | 6,56 | 1,20 | 0,00 |
| 30.01.2024 | 4 | CEN/EX1 | 0,88 | 0,14 | 0,00 |
| 01.02.2024 | 4 | CEN/EX1 | 2,31 | 2,87 | 0,13 |
| 06.02.2024 | 4 | CEN/EX1 | 2,33 | 1,33 | 0,00 |
| 25.01.2024 | 4 | CEN/EX2 | 5,73 | 1,71 | 0,00 |
| 30.01.2024 | 4 | CEN/EX2 | 10,13 | 1,01 | 0,00 |
| 01.02.2024 | 4 | CEN/EX2 | 8,38 | 1,13 | 0,28 |
| 06.02.2024 | 4 | CEN/EX2 | 5,62 | 0,75 | 0,15 |

Table 3. Raw data of spike experiment to determine the recovery efficiency of influenza A and B virus

| Sample description | sample group | Target | Gene copies / µl RNA extract | Gene copies / l waste water |
| --- | --- | --- | --- | --- |
| N1.1 | Negative | IAV | 0,00 | 0,00 |
| N3.1 | Negative | IAV | 0,00 | 0,00 |
| N3.1 | Negative | IBV | 0,00 | 0,00 |
| N1.2 | Negative | IAV | 0,00 | 0,00 |
| N3.2 | Negative | IAV | 0,00 | 0,00 |
| N1.3 | Negative | IAV | 0,00 | 0,00 |
| N3.3 | Negative | IAV | 0,00 | 0,00 |
| N3.3 | Negative | IBV | 0,00 | 0,00 |
| N1.4 | Negative | IAV | 0,00 | 0,00 |
| N1.4 | Negative | IBV | 0,00 | 0,00 |
| N3.4 | Negative | IAV | 0,00 | 0,00 |
| N3.4 | Negative | IBV | 0,00 | 0,00 |
| N2.1 | Negative | IAV | 0,00 | 0,00 |
| N2.1 | Negative | IBV | 0,00 | 0,00 |
| N2.2 | Negative | IAV | 0,00 | 0,00 |
| N2.2 | Negative | IBV | 0,00 | 0,00 |
| N2.3 | Negative | IAV | 0,00 | 0,00 |
| N2.4 | Negative | IAV | 0,00 | 0,00 |
| N2.4 | Negative | IBV | 0,00 | 0,00 |
| N2.3 | Negative | IBV | 0,14 | 676,64 |
| N1.1 | Negative | IBV | 0,15 | 734,57 |
| N1.2 | Negative | IBV | 0,15 | 735,69 |
| N3.2 | Negative | IBV | 0,15 | 748,89 |
| N1.3 | Negative | IBV | 0,16 | 775,53 |
| G2.4 | 10.000 [gc/l] | IAV | 0,46 | 2288,00 |
| G1.2 | 10.000 [gc/l] | IAV | 0,88 | 4390,90 |
| G1.2 | 10.000 [gc/l] | IBV | 0,88 | 4390,90 |
| G2.4 | 10.000 [gc/l] | IBV | 0,92 | 4576,37 |
| G2.3 | 10.000 [gc/l] | IBV | 1,18 | 5891,55 |
| G3.1 | 10.000 [gc/l] | IBV | 1,18 | 5907,43 |
| G1.3 | 10.000 [gc/l] | IBV | 1,19 | 5940,07 |
| G3.4 | 10.000 [gc/l] | IAV | 1,28 | 6386,54 |
| G3.3 | 10.000 [gc/l] | IAV | 1,29 | 6447,43 |
| G1.1 | 10.000 [gc/l] | IAV | 1,35 | 6748,00 |
| G1.1 | 10.000 [gc/l] | IBV | 1,35 | 6748,00 |
| G2.1 | 10.000 [gc/l] | IAV | 1,46 | 7319,75 |
| G3.1 | 10.000 [gc/l] | IAV | 1,48 | 7384,68 |
| G1.3 | 10.000 [gc/l] | IAV | 1,49 | 7425,48 |
| G3.2 | 10.000 [gc/l] | IBV | 1,58 | 7881,81 |
| G2.1 | 10.000 [gc/l] | IBV | 1,63 | 8133,29 |
| G2.2 | 10.000 [gc/l] | IBV | 1,68 | 8387,04 |
| G3.3 | 10.000 [gc/l] | IBV | 1,72 | 8597,24 |
| G1.4 | 10.000 [gc/l] | IBV | 1,80 | 8987,94 |
| G1.4 | 10.000 [gc/l] | IAV | 2,25 | 11235,82 |
| G3.4 | 10.000 [gc/l] | IBV | 2,27 | 11355,87 |
| G2.2 | 10.000 [gc/l] | IAV | 2,29 | 11438,12 |
| G2.3 | 10.000 [gc/l] | IAV | 2,36 | 11785,59 |
| G3.2 | 10.000 [gc/l] | IAV | 2,72 | 13616,83 |
| M1.2 | 100.000 [gc/l] | IBV | 7,41 | 37048,78 |
| M1.2 | 100.000 [gc/l] | IAV | 8,70 | 43502,08 |
| M3.1 | 100.000 [gc/l] | IAV | 10,34 | 51693,90 |
| M1.4 | 100.000 [gc/l] | IBV | 10,99 | 54955,80 |
| M3.3 | 100.000 [gc/l] | IBV | 12,23 | 61131,43 |
| M1.1 | 100.000 [gc/l] | IAV | 12,24 | 61207,88 |
| M3.3 | 100.000 [gc/l] | IAV | 12,51 | 62539,91 |
| M3.4 | 100.000 [gc/l] | IAV | 12,65 | 63253,74 |
| M1.4 | 100.000 [gc/l] | IAV | 12,99 | 64971,01 |
| M3.4 | 100.000 [gc/l] | IBV | 13,07 | 65367,13 |
| M2.4 | 100.000 [gc/l] | IBV | 13,23 | 66157,47 |
| M2.4 | 100.000 [gc/l] | IAV | 13,43 | 67162,26 |
| M1.1 | 100.000 [gc/l] | IBV | 13,50 | 67482,91 |
| M3.1 | 100.000 [gc/l] | IBV | 13,59 | 67935,90 |
| M2.3 | 100.000 [gc/l] | IBV | 13,80 | 68997,09 |
| M2.1 | 100.000 [gc/l] | IAV | 15,33 | 76664,01 |
| M3.2 | 100.000 [gc/l] | IBV | 15,64 | 78213,38 |
| M1.3 | 100.000 [gc/l] | IBV | 15,65 | 78248,37 |
| M2.2 | 100.000 [gc/l] | IBV | 16,12 | 80576,22 |
| M2.1 | 100.000 [gc/l] | IBV | 16,33 | 81647,52 |
| M2.3 | 100.000 [gc/l] | IAV | 17,15 | 85752,99 |
| M1.3 | 100.000 [gc/l] | IAV | 17,74 | 88714,66 |
| M2.2 | 100.000 [gc/l] | IAV | 18,34 | 91701,15 |
| M3.2 | 100.000 [gc/l] | IAV | 19,76 | 98788,54 |
| H1.4 | 1.000.000 [gc/l] | IBV | 48,22 | 241122,03 |
| H1.4 | 1.000.000 [gc/l] | IAV | 50,28 | 251402,45 |
| H1.3 | 1.000.000 [gc/l] | IAV | 123,52 | 617622,67 |
| H2.4 | 1.000.000 [gc/l] | IBV | 125,88 | 629392,37 |
| H1.3 | 1.000.000 [gc/l] | IBV | 128,67 | 643328,14 |
| H2.4 | 1.000.000 [gc/l] | IAV | 138,37 | 691832,90 |
| H2.3 | 1.000.000 [gc/l] | IBV | 139,17 | 695838,67 |
| H3.2 | 1.000.000 [gc/l] | IBV | 141,33 | 706640,92 |
| H2.1 | 1.000.000 [gc/l] | IBV | 141,56 | 707808,98 |
| H3.3 | 1.000.000 [gc/l] | IBV | 141,84 | 709199,82 |
| H1.1 | 1.000.000 [gc/l] | IBV | 142,15 | 710771,52 |
| H3.3 | 1.000.000 [gc/l] | IAV | 144,74 | 723693,00 |
| H1.1 | 1.000.000 [gc/l] | IAV | 144,91 | 724535,96 |
| H2.1 | 1.000.000 [gc/l] | IAV | 145,43 | 727163,53 |
| H2.3 | 1.000.000 [gc/l] | IAV | 145,66 | 728305,99 |
| H3.4 | 1.000.000 [gc/l] | IBV | 152,50 | 762514,67 |
| H3.1 | 1.000.000 [gc/l] | IAV | 155,75 | 778762,39 |
| H3.2 | 1.000.000 [gc/l] | IAV | 156,38 | 781904,43 |
| H2.2 | 1.000.000 [gc/l] | IBV | 161,79 | 808941,14 |
| H3.1 | 1.000.000 [gc/l] | IBV | 164,14 | 820705,41 |
| H3.4 | 1.000.000 [gc/l] | IAV | 164,56 | 822800,53 |
| H2.2 | 1.000.000 [gc/l] | IAV | 169,02 | 845122,27 |

Table 4. Raw data of IAV, IBV, and PMMoV Across the Winter Season 2023/2024 across four WWTPs located in Mecklenburg Western Pomerania, Germany.

| **Date** | **WTTP** | **PMMoV** | **IAV** | **IBV** |
| --- | --- | --- | --- | --- |
|  |  | **[gene copies / liter wastewater]** | | |
| 14.11.2023 | 1 | 2,60E+08 | 1,26E+03 | 8,40E+02 |
| 14.11.2023 | 2 | 2,37E+08 | 0,00E+00 | 0,00E+00 |
| 14.11.2023 | 3 | 2,34E+08 | 0,00E+00 | 0,00E+00 |
| 14.11.2023 | 4 | 3,58E+06 | 2,13E+03 | 0,00E+00 |
| 16.11.2023 | 1 | 2,90E+08 | 0,00E+00 | 6,18E+02 |
| 16.11.2023 | 4 | 2,67E+08 | 6,31E+02 | 0,00E+00 |
| 16.11.2023 | 2 | 3,06E+06 | 3,17E+02 | 0,00E+00 |
| 16.11.2023 | 3 | 5,91E+08 | 1,33E+03 | 0,00E+00 |
| 21.11.2023 | 1 | 5,91E+08 | 6,44E+02 | 0,00E+00 |
| 21.11.2023 | 4 | 6,44E+08 | 9,07E+02 | 0,00E+00 |
| 21.11.2023 | 2 | 7,11E+07 | 0,00E+00 | 0,00E+00 |
| 21.11.2023 | 3 | 6,67E+08 | 1,77E+03 | 0,00E+00 |
| 23.11.2023 | 2 | 3,33E+08 | 3,04E+02 | 0,00E+00 |
| 23.11.2023 | 4 | 3,83E+08 | 0,00E+00 | 0,00E+00 |
| 23.11.2023 | 1 | 4,93E+08 | 0,00E+00 | 0,00E+00 |
| 23.11.2023 | 3 | 5,20E+08 | 0,00E+00 | 0,00E+00 |
| 28.11.2023 | 2 | 1,16E+08 | 0,00E+00 | 0,00E+00 |
| 28.11.2023 | 4 | 9,33E+07 | 6,31E+02 | 0,00E+00 |
| 28.11.2023 | 1 | 6,84E+07 | 4,41E+02 | 0,00E+00 |
| 28.11.2023 | 3 | 8,53E+07 | 6,71E+03 | 0,00E+00 |
| 30.11.2023 | 1 | 2,23E+08 | 0,00E+00 | 0,00E+00 |
| 30.11.2023 | 3 | 1,80E+08 | 5,78E+02 | 0,00E+00 |
| 30.11.2023 | 4 | 1,49E+08 | 2,95E+02 | 2,95E+02 |
| 30.11.2023 | 2 | 2,19E+09 | 0,00E+00 | 0,00E+00 |
| 05.12.2023 | 4 | 2,16E+08 | 2,01E+03 | 4,02E+02 |
| 05.12.2023 | 1 | 3,03E+08 | 4,26E+03 | 3,28E+02 |
| 05.12.2023 | 2 | 2,54E+08 | 1,64E+03 | 0,00E+00 |
| 05.12.2023 | 3 | 2,84E+08 | 8,98E+03 | 0,00E+00 |
| 07.12.2023 | 1 | 2,67E+08 | 3,57E+03 | 2,38E+03 |
| 07.12.2023 | 2 | 2,46E+08 | 8,44E+02 | 0,00E+00 |
| 07.12.2023 | 3 | 3,73E+08 | 1,49E+04 | 0,00E+00 |
| 07.12.2023 | 4 | 3,07E+08 | 2,10E+03 | 3,50E+02 |
| 12.12.2023 | 1 | 1,1E+08 | 2,8E+03 | 3,5E+03 |
| 12.12.2023 | 4 | 1,0E+08 | 2,1E+03 | 0,0E+00 |
| 12.12.2023 | 2 | 1,8E+08 | 1,5E+03 | 0,0E+00 |
| 12.12.2023 | 3 | 3,3E+08 | 1,9E+04 | 0,0E+00 |
| 14.12.2023 | 1 | 1,6E+08 | 2,2E+04 | 6,5E+03 |
| 14.12.2023 | 2 | 2,2E+08 | 5,9E+03 | 0,0E+00 |
| 14.12.2023 | 3 | 2,1E+08 | 4,1E+04 | 0,0E+00 |
| 14.12.2023 | 4 | 1,8E+08 | 6,8E+03 | 0,0E+00 |
| 19.12.2023 | 1 | 0,0E+00 | 7,7E+03 | 1,9E+03 |
| 19.12.2023 | 2 | 0,0E+00 | 4,9E+03 | 1,1E+03 |
| 19.12.2023 | 3 | 0,0E+00 | 4,0E+04 | 0,0E+00 |
| 19.12.2023 | 4 | 0,0E+00 | 1,9E+04 | 1,3E+03 |
| 21.12.2023 | 1 | 1,6E+08 | 5,0E+04 | 1,3E+04 |
| 21.12.2023 | 2 | 1,6E+08 | 9,7E+03 | 0,0E+00 |
| 21.12.2023 | 4 | 1,2E+08 | 1,4E+04 | 4,1E+03 |
| 21.12.2023 | 3 | 1,8E+08 | 7,8E+04 | 0,0E+00 |
| 26.12.2023 | 2 | 1,1E+08 | 1,7E+04 | 7,5E+02 |
| 28.12.2023 | 2 | 1,1E+08 | 7,0E+03 | 0,0E+00 |
| 02.01.2024 | 2 | 1,6E+08 | 1,5E+04 | 6,8E+02 |
| 04.01.2024 | 2 | 1,3E+08 | 1,1E+04 | 3,6E+02 |
| 04.01.2024 | 1 | 1,3E+08 | 1,1E+04 | 0,0E+00 |
| 04.01.2024 | 4 | 1,3E+08 | 8,7E+03 | 0,0E+00 |
| 04.01.2024 | 3 | 1,8E+08 | 4,8E+04 | 4,1E+02 |
| 09.01.2024 | 1 | 1,3E+08 | 1,5E+04 | 1,3E+04 |
| 09.01.2024 | 2 | 6,0E+07 | 1,7E+04 | 1,6E+03 |
| 09.01.2024 | 3 | 1,6E+08 | 7,7E+04 | 2,0E+03 |
| 09.01.2024 | 4 | 1,8E+08 | 7,0E+03 | 3,9E+02 |
| 11.01.2024 | 1 | 1,1E+08 | 2,6E+04 | 9,0E+03 |
| 11.01.2024 | 2 | 9,3E+07 | 2,8E+04 | 3,9E+03 |
| 11.01.2024 | 3 | 6,4E+07 | 3,4E+04 | 6,7E+02 |
| 11.01.2024 | 4 | 1,3E+08 | 1,7E+04 | 0,0E+00 |
| 16.01.2024 | 1 | 3,2E+08 | 3,4E+04 | 1,7E+04 |
| 16.01.2024 | 2 | 3,2E+08 | 2,1E+04 | 7,5E+03 |
| 16.01.2024 | 3 | 3,7E+08 | 7,7E+04 | 0,0E+00 |
| 16.01.2024 | 4 | 2,5E+08 | 3,8E+04 | 6,9E+02 |
| 18.01.2024 | 2 | 1,9E+08 | 4,0E+04 | 1,5E+04 |
| 18.01.2024 | 1 | 2,3E+08 | 3,2E+04 | 2,5E+04 |
| 18.01.2024 | 4 | 2,9E+08 | 2,4E+04 | 0,0E+00 |
| 18.01.2024 | 3 | 3,7E+08 | 4,3E+04 | 0,0E+00 |
| 23.01.2024 | 1 | 1,5E+08 | 6,3E+04 | 1,1E+04 |
| 23.01.2024 | 2 | 1,5E+08 | 2,8E+04 | 1,2E+04 |
| 23.01.2024 | 3 | 1,4E+08 | 4,7E+04 | 6,4E+02 |
| 23.01.2024 | 4 | 1,6E+08 | 3,2E+04 | 0,0E+00 |
| 25.01.2024 | 1 | 1,4E+08 | 7,2E+04 | 1,0E+04 |
| 25.01.2024 | 2 | 9,8E+07 | 4,6E+04 | 1,6E+04 |
| 25.01.2024 | 3 | 1,4E+08 | 9,0E+04 | 6,9E+02 |
| 25.01.2024 | 4 | 1,3E+08 | 8,0E+04 | 2,5E+03 |
| 30.01.2024 | 1 | 2,1E+08 | 2,2E+05 | 2,9E+04 |
| 30.01.2024 | 2 | 1,9E+08 | 6,6E+04 | 1,4E+04 |
| 30.01.2024 | 3 | 1,8E+08 | 1,1E+05 | 6,7E+02 |
| 30.01.2024 | 4 | 2,7E+08 | 7,8E+04 | 6,5E+02 |
| 01.02.2024 | 1 | 1,3E+08 | 7,6E+04 | 9,5E+03 |
| 01.02.2024 | 2 | 2,1E+08 | 6,1E+04 | 2,2E+04 |
| 01.02.2024 | 3 | 1,4E+08 | 1,1E+05 | 2,0E+03 |
| 01.02.2024 | 4 | 1,7E+08 | 4,5E+04 | 8,4E+03 |
| 06.02.2024 | 1 | 5,8E+07 | 4,6E+04 | 1,1E+04 |
| 06.02.2024 | 2 | 1,0E+08 | 3,2E+04 | 2,9E+04 |
| 06.02.2024 | 3 | 8,2E+07 | 1,0E+05 | 2,6E+04 |
| 06.02.2024 | 4 | 7,2E+07 | 7,1E+04 | 1,3E+03 |
| 08.02.2024 | 1 | 1,5E+08 | 6,7E+04 | 1,4E+04 |
| 08.02.2024 | 2 | 1,4E+08 | 4,5E+04 | 4,5E+04 |
| 08.02.2024 | 3 | 2,3E+08 | 8,8E+04 | 2,0E+04 |
| 08.02.2024 | 4 | 2,7E+08 | 9,5E+04 | 1,3E+03 |
| 13.02.2024 | 1 | 6,5E+07 | 1,1E+04 | 1,5E+04 |
| 13.02.2024 | 2 | 5,4E+07 | 1,2E+04 | 2,5E+04 |
| 13.02.2024 | 3 | 9,8E+07 | 5,3E+04 | 1,4E+04 |
| 13.02.2024 | 4 | 9,8E+07 | 6,9E+04 | 2,4E+03 |
| 15.02.2024 | 1 | 1,1E+08 | 4,8E+04 | 2,0E+04 |
| 15.02.2024 | 2 | 1,1E+08 | 1,5E+04 | 3,5E+04 |
| 15.02.2024 | 3 | 1,4E+08 | 5,5E+04 | 1,3E+04 |
| 15.02.2024 | 4 | 1,0E+08 | 9,2E+04 | 1,3E+03 |
| 20.02.2024 | 1 | 1,4E+08 | 1,7E+04 | 5,8E+03 |
| 20.02.2024 | 2 | 1,0E+08 | 3,5E+04 | 2,4E+04 |
| 20.02.2024 | 3 | 1,9E+08 | 3,8E+04 | 8,0E+03 |
| 20.02.2024 | 4 | 1,5E+08 | 5,9E+04 | 0,0E+00 |
| 22.02.2024 | 1 | 7,2E+07 | 1,1E+04 | 1,0E+04 |
| 22.02.2024 | 2 | 6,7E+07 | 1,7E+04 | 4,6E+04 |
| 22.02.2024 | 3 | 1,2E+08 | 3,1E+04 | 2,9E+04 |
| 22.02.2024 | 4 | 1,1E+08 | 9,1E+04 | 7,3E+02 |
| 27.02.2024 | 1 | 6,6E+07 | 1,9E+04 | 8,3E+03 |
| 27.02.2024 | 2 | 6,0E+07 | 6,2E+03 | 1,1E+04 |
| 27.02.2024 | 3 | 2,6E+06 | 2,4E+04 | 1,2E+04 |
| 27.02.2024 | 4 | 1,3E+08 | 4,8E+04 | 2,7E+03 |
| 29.02.2024 | 1 | 8,8E+07 | 1,9E+04 | 5,8E+03 |
| 29.02.2024 | 2 | 8,0E+07 | 1,1E+04 | 1,4E+04 |
| 29.02.2024 | 3 | 1,5E+08 | 3,4E+04 | 7,1E+03 |
| 29.02.2024 | 4 | 1,7E+08 | 6,8E+04 | 7,2E+02 |
| 05.03.2024 | 1 | 1,1E+08 | 1,1E+04 | 5,3E+03 |
| 05.03.2024 | 2 | 5,7E+07 | 8,4E+03 | 1,1E+04 |
| 05.03.2024 | 3 | 1,1E+08 | 3,3E+04 | 3,8E+04 |
| 05.03.2024 | 4 | 2,2E+08 | 4,2E+04 | 4,7E+03 |
| 07.03.2024 | 1 | 1,4E+08 | 5,9E+03 | 4,5E+03 |
| 07.03.2024 | 2 | 9,9E+07 | 6,2E+03 | 1,1E+04 |
| 07.03.2024 | 3 | 1,5E+08 | 2,7E+04 | 1,6E+04 |
| 07.03.2024 | 4 | 2,6E+08 | 3,2E+04 | 7,5E+02 |
| 12.03.2024 | 1 | 1,9E+08 | 9,3E+03 | 6,6E+03 |
| 12.03.2024 | 2 | 2,0E+07 | 1,1E+04 | 3,2E+04 |
| 12.03.2024 | 3 | 2,1E+08 | 8,4E+03 | 9,1E+03 |
| 12.03.2024 | 4 | 3,0E+08 | 1,1E+04 | 8,6E+03 |
| 14.03.2024 | 1 | 3,2E+08 | 6,2E+03 | 6,9E+03 |
| 14.03.2024 | 2 | 3,2E+08 | 4,0E+03 | 2,3E+04 |
| 14.03.2024 | 3 | 5,6E+08 | 9,3E+03 | 2,1E+04 |
| 14.03.2024 | 4 | 6,9E+08 | 1,2E+04 | 1,0E+04 |
| 19.03.2024 | 4 | 3,5E+08 | 2,7E+03 | 0,0E+00 |
| 19.03.2024 | 3 | 1,7E+08 | 3,8E+03 | 2,2E+04 |
| 19.03.2024 | 2 | 1,3E+08 | 6,4E+03 | 2,1E+04 |
| 19.03.2024 | 1 | 1,3E+08 | 3,8E+03 | 4,5E+02 |
| 21.03.2024 | 4 | 1,6E+08 | 8,7E+03 | 1,3E+04 |
| 21.03.2024 | 3 | 3,5E+08 | 6,9E+02 | 2,1E+04 |
| 21.03.2024 | 2 | 1,1E+08 | 6,9E+03 | 1,4E+04 |
| 21.03.2024 | 1 | 1,1E+08 | 6,3E+03 | 8,9E+03 |
| 26.03.2024 | 1 | 7,8E+08 | 2,0E+03 | 1,4E+04 |
| 26.03.2024 | 2 | 8,6E+08 | 1,7E+04 | 2,3E+04 |
| 26.03.2024 | 3 | 4,0E+08 | 2,4E+04 | 1,7E+04 |
| 26.03.2024 | 4 | 4,6E+08 | 4,8E+03 | 2,2E+04 |
| 28.03.2024 | 2 | 1,1E+08 | 4,5E+04 | 1,5E+04 |
| 02.04.2024 | 1 | 1,7E+08 | 1,9E+03 | 1,5E+04 |
| 02.04.2024 | 2 | 1,3E+08 | 5,6E+03 | 2,3E+04 |
| 02.04.2024 | 3 | 3,9E+08 | 5,2E+03 | 2,7E+04 |
| 02.04.2024 | 4 | 2,0E+08 | 2,5E+03 | 1,0E+04 |
| 04.04.2024 | 1 | 1,8E+08 | 2,6E+03 | 1,3E+04 |
| 04.04.2024 | 2 | 1,4E+08 | 6,6E+03 | 1,1E+04 |
| 04.04.2024 | 3 | 3,5E+08 | 8,1E+03 | 2,3E+04 |
| 04.04.2024 | 4 | 2,1E+08 | 1,3E+03 | 1,0E+04 |
| 09.04.2024 | 1 | 1,3E+08 | 6,4E+02 | 1,2E+04 |
| 09.04.2024 | 2 | 1,3E+08 | 1,3E+03 | 3,8E+03 |
| 09.04.2024 | 3 | 1,7E+08 | 6,0E+02 | 1,6E+04 |
| 09.04.2024 | 4 | 1,9E+08 | 2,0E+03 | 2,7E+03 |
| 11.04.2024 | 1 | 1,8E+08 | 6,5E+02 | 4,6E+03 |
| 11.04.2024 | 2 | 1,0E+08 | 2,0E+03 | 5,2E+03 |
| 11.04.2024 | 3 | 1,9E+08 | 2,0E+03 | 1,4E+04 |
| 11.04.2024 | 4 | 1,6E+08 | 2,0E+03 | 2,7E+03 |
| 16.04.2024 | 1 | 1,9E+08 | 0,0E+00 | 7,3E+03 |
| 16.04.2024 | 2 | 2,6E+08 | 4,8E+02 | 4,8E+02 |
| 16.04.2024 | 3 | 2,6E+08 | 1,7E+03 | 6,2E+03 |
| 16.04.2024 | 4 | 2,4E+08 | 0,0E+00 | 2,7E+03 |
| 18.04.2024 | 1 | 1,4E+08 | 3,2E+02 | 3,2E+03 |
| 18.04.2024 | 2 | 1,1E+08 | 3,6E+02 | 3,6E+02 |
| 18.04.2024 | 3 | 2,0E+08 | 7,0E+02 | 1,8E+03 |
| 18.04.2024 | 4 | 2,8E+08 | 7,0E+02 | 2,4E+03 |
| 23.04.2024 | 1 | 1,1E+08 | 0,0E+00 | 0,0E+00 |
| 23.04.2024 | 2 | 1,9E+08 | 6,6E+02 | 6,6E+02 |
| 23.04.2024 | 3 | 3,1E+08 | 0,0E+00 | 3,9E+03 |
| 23.04.2024 | 4 | 3,1E+08 | 2,1E+03 | 4,2E+03 |
| 25.04.2024 | 1 | 1,4E+08 | 7,0E+02 | 2,8E+03 |
| 25.04.2024 | 2 | 9,8E+07 | 6,9E+02 | 1,4E+03 |
| 25.04.2024 | 3 | 1,8E+08 | 6,7E+02 | 4,7E+03 |
| 25.04.2024 | 4 | 2,0E+08 | 3,3E+03 | 6,7E+02 |
| 30.04.2024 | 1 | 2,5E+08 | 6,6E+02 | 5,3E+03 |
| 30.04.2024 | 2 | 2,1E+08 | 2,5E+03 | 6,3E+02 |
| 30.04.2024 | 3 | 2,4E+08 | 0,0E+00 | 4,6E+03 |
| 30.04.2024 | 4 | 1,9E+08 | 2,1E+03 | 0,0E+00 |
| 02.05.2024 | 1 | 2,1E+08 | 0,0E+00 | 7,2E+03 |
| 02.05.2024 | 2 | 2,0E+08 | 2,5E+03 | 2,5E+03 |
| 02.05.2024 | 4 | 2,2E+08 | 0,0E+00 | 1,3E+03 |
| 07.05.2024 | 1 | 2,0E+08 | 0,0E+00 | 2,7E+03 |
| 07.05.2024 | 2 | 1,5E+08 | 0,0E+00 | 0,0E+00 |
| 07.05.2024 | 3 | 3,5E+08 | 6,8E+02 | 4,1E+03 |
| 07.05.2024 | 4 | 1,9E+08 | 7,0E+02 | 0,0E+00 |
| 14.05.2024 | 1 | 1,6E+08 | 0,0E+00 | 2,0E+03 |
| 14.05.2024 | 2 | 8,3E+07 | 0,0E+00 | 0,0E+00 |
| 14.05.2024 | 3 | 2,1E+08 | 0,0E+00 | 1,7E+03 |
| 14.05.2024 | 4 | 4,0E+08 | 0,0E+00 | 1,5E+03 |
| 09.05.2024 | 1 | 1,8E+08 | 0,0E+00 | 1,4E+03 |
| 16.05.2024 | 1 | 1,5E+08 | 0,0E+00 | 2,6E+03 |
| 16.05.2024 | 2 | 1,7E+08 | 1,4E+03 | 0,0E+00 |
| 16.05.2024 | 3 | 2,7E+08 | 7,7E+02 | 7,7E+02 |
| 16.05.2024 | 4 | 2,7E+08 | 0,0E+00 | 1,8E+03 |
| 21.05.2024 | 1 | 9,0E+07 | 0,0E+00 | 2,7E+03 |
| 21.05.2024 | 2 | 2,9E+08 | 0,0E+00 | 1,3E+03 |
| 21.05.2024 | 4 | 1,3E+08 | 0,0E+00 | 6,0E+02 |
| 23.05.2024 | 1 | 1,0E+08 | 0,0E+00 | 7,0E+02 |
| 23.05.2024 | 2 | 1,4E+08 | 0,0E+00 | 0,0E+00 |
| 23.05.2024 | 3 | 2,3E+08 | 0,0E+00 | 6,0E+02 |
| 23.05.2024 | 4 | 1,2E+08 | 0,0E+00 | 2,1E+03 |
| 28.05.2024 | 1 | 1,7E+08 | 0,0E+00 | 0,0E+00 |
| 28.05.2024 | 2 | 1,9E+08 | 0,0E+00 | 6,0E+02 |
| 28.05.2024 | 3 | 2,4E+08 | 0,0E+00 | 0,0E+00 |
| 28.05.2024 | 4 | 1,5E+08 | 0,0E+00 | 0,0E+00 |
| 30.05.2024 | 1 | 1,3E+08 | 0,0E+00 | 0,0E+00 |
| 30.05.2024 | 2 | 3,1E+08 | 0,0E+00 | 0,0E+00 |
| 30.05.2024 | 3 | 2,3E+08 | 0,0E+00 | 0,0E+00 |
| 30.05.2024 | 4 | 7,5E+08 | 0,0E+00 | 1,3E+03 |
